# Fertility in the Shadow of Cancer: Experiences of Reproductive Loss Among Women with Gynecological Cancers in Ghana

**DOI:** 10.64898/2026.02.21.26346234

**Authors:** Agani Afaya, Donald Babamomba Amenah, Fahd Chambas, Promise Aidoo, Ohene Addo Gideon, Mavis Vidzor, Bernice Aidoo, Richard Adongo Afaya, Mabel Apaanye Avane, Silas Selorm Daniels-Donkor, Dennis Bomansang Daliri, Solomon Mohammed Salia

## Abstract

**Background:** Gynecological cancers and their treatments can compromise fertility, with profound psychosocial consequences for women of reproductive age. Yet, women’s lived experiences of cancer-related infertility remain underexplored in low-resource settings, including Ghana. This study examined the impact of gynecological cancers on fertility among reproductive-aged women receiving care at Ho Teaching Hospital, Ghana.

**Methods:** A qualitative descriptive design was used. Fourteen women aged 15–49 years with gynecological cancers who had completed or were undergoing treatment were purposively recruited until saturation. Semi-structured interviews (30–45 minutes) were conducted face-to-face or by telephone in English, Twi, or Ewe, audio-recorded, transcribed verbatim, and analyzed using thematic analysis. Strategies to enhance rigor included independent coding, member checking, reflexivity, and peer debriefing.

**Results:** Five themes and eighteen subthemes emerged. Participants described infertility as a threat to womanhood and future life plans, expressed as a sense of incompleteness, fear of rejection, denial, and shattered aspirations. Social consequences included stigma and impaired intimate relationships. Treatment-related burdens, menstrual changes, pain, fatigue, and anxiety compounded distress. Economic hardship and educational disruption were common. Women also demonstrated resilience through adherence to treatment, dietary and lifestyle modifications, faith-based coping, and family support.

**Conclusion:** Gynecological cancer–related infertility is a multidimensional survivorship burden. Integrating fertility counseling, psychosocial support, symptom management, and financial/social protection into cancer care is critical in Ghanaian settings.

## Background

Gynecological cancers remain a major yet often under-recognized threat to women’s health worldwide, exacting a profound toll on survival, quality of life, and health systems, particularly in low- and middle-income settings (1). In 2022, approximately 1.47 million women were newly diagnosed with gynecological cancers, with an estimated 680,000 deaths globally, representing over 15% of all cancer diagnoses and cancer-related deaths among women (2,3). The burden is projected to increase markedly by 2050, with the steepest rises in incidence and mortality anticipated in low- and middle-income regions, particularly sub-Saharan Africa (1). In Ghana, gynaecological cancers, particularly cervical and ovarian cancers, constitute a major source of cancer-related morbidity and mortality among women, with cervical cancer alone accounting for an estimated 2,797 new cases and 1,699 deaths annually, corresponding to age-standardised incidence and mortality rates of approximately 27.0 and 16.9 per 100,000 women, respectively (4,5). These outcomes are frequently shaped by late-stage diagnosis, low screening uptake, estimated at about 7% among eligible women, and constrained access to specialised oncology services, resulting in substantial out-of-pocket treatment costs and poorer survival outcomes (6,7). Within this context, the consequences of gynecological cancers extend beyond survival to profoundly affect women’s reproductive health, social roles, and economic stability, particularly for those diagnosed during their reproductive years.

Gynecological cancers comprise a heterogeneous group of malignancies, including cervical, vulvar, ovarian, vaginal, uterine, and fallopian tube. Among these, fallopian tube tumors are the rarest (8), while endometrial, cervical, and ovarian cancers are the most prevalent, together accounting for more than one-third of newly diagnosed cancers among women worldwide (2,4,9,10). Advances in oncological management, including chemotherapy, surgery, pelvic or abdominal radiotherapy, and hormonotherapy, have improved survival outcomes; however, these treatments are frequently associated with adverse reproductive consequences such as infertility, premature ovarian failure, and early menopause, resulting in the loss of reproductive potential (11,12). Consequently, as the population of gynecological cancer survivors continues to grow, concerns related to fertility preservation and pregnancy outcomes have become increasingly salient for women of reproductive age (13).

Gynecological cancers are influenced by hormonal and reproductive factors, suggesting potential intersections with female infertility. However, whether infertility itself is associated with an increased risk of developing gynecological cancers remains a subject of ongoing debate (14). Growing global rates of female infertility have further intensified interest in this potential relationship (14,15). Although several epidemiological studies have examined associations between female infertility and gynecological cancers, findings remain inconsistent and, at times, contradictory (14–18). Importantly, existing research has largely focused on biomedical or epidemiological associations, with limited attention to women’s lived experiences of infertility following gynecological cancer diagnosis and treatment.

Given the rising global burden of gynecological cancers and the fertility-compromising nature of their treatment, infertility represents not only a clinical outcome but also a profound psychosocial concern for women of reproductive age who desire future childbearing. Understanding women’s experiences in this context is particularly critical in low-resource settings, where access to fertility preservation and psychosocial support may be limited. Therefore, this study explored the impact of gynecological cancers and their treatment on fertility among women of reproductive age receiving care at Ho Teaching Hospital in Ghana.

## Methods

### Study Design

This study adopted a qualitative descriptive design to explore the impact of gynecological cancer and its treatments on fertility among women of reproductive age. The study was reported in line with the Consolidated criteria for reporting qualitative research (COREQ) (19).

### Study Setting

The study was conducted at the Ho Teaching Hospital (HTH), a tertiary referral facility in the Volta Region of Ghana. The hospital serves as the main teaching and research hospital affiliated with the University of Health and Allied Sciences (UHAS), providing specialized services in various disciplines, including obstetrics and gynecology. The gynecology unit offers both routine and specialized care, including the management of gynecological cancers through surgical interventions, chemotherapy, and palliative care means, while also engaging in preventive measures such as screening and community education. This makes the hospital a critical setting for examining issues related to women’s reproductive health and cancer care.

### Study Population

The target population consisted of reproductive-aged women (15–49 years) who had been diagnosed with and treated for gynecological cancers, including but not limited to cervical, ovarian, uterine, and endometrial cancers.

### Inclusion and Exclusion Criteria

The inclusion criteria for participants in this study were as follows:

- Women aged 18–49 years.
- Women diagnosed with any type of gynecological cancer.
- Women who have completed or are currently undergoing treatment (surgery, chemotherapy, and/or radiotherapy).
- Willingness to participate and provide informed consent.

The exclusion criteria involved:

- Minors (that is, adolescents under 18) were excluded.
- Women diagnosed with non-gynecological cancers.
- Women with known infertility prior to cancer diagnosis.
- Women with mental health issues or critically ill.

### Sampling Strategy

This study employed a purposive sampling strategy to recruit participants who met the inclusion criteria and had firsthand experience relevant to the study objectives. Purposive sampling was selected because it allows for the intentional identification of information-rich participants who can provide in-depth and meaningful insights into the phenomenon under investigation (20). After establishing the eligibility criteria, the researchers used their judgment and familiarity with the study context to identify and approach potential participants who met these requirements (21).

### Data Collection Method and Procedure

A semi-structured interview guide was developed based on insights from the literature review (22) and aligned with the study’s objectives. The guide was written in English and organized into sections that reflect the key objectives of the study. It included both open-ended questions and probing prompts to encourage participants to elaborate on their responses and share in-depth experiences. The guide was pre-tested with three participants of similar characteristics to ensure clarity, cultural appropriateness, and relevance before being finalized for use in the main study.

Data were collected through semi-structured interviews. Participant recruitment occurred between 01/11/2025 and 14/11/2025. Eligible individuals were contacted, informed about the purpose of the study, and provided with participant information sheets. Written informed consent was obtained before participation. Interviews were primarily conducted face-to-face in private and convenient locations to ensure confidentiality and comfort. However, for participants who were unable to attend in person due to distance, time constraints, or health-related reasons, telephone interviews were conducted as an alternative. Each interview session lasted between 30 and 45 minutes and was conducted in English, Twi, or Ewe, depending on participants’ fluency. Interviews in Twi and Ewe were translated into English by a multilingual team member. To protect meaning, a second multilingual team member verified the translations and resolved any inconsistencies through discussion. With participants’ permission, all interviews were audio-recorded, and a reflexive journal was maintained throughout data collection to document non-verbal cues observed during face-to-face interviews as well as capture the researcher’s reflections and analytic insights. Probing questions were used to encourage elaboration and ensure rich narratives. Sampling and interviewing were conducted iteratively in conjunction with preliminary analysis. Data saturation was judged by determining if new interviews generated additional codes or influenced existing themes. Saturation was determined to be reached when successive interviews produced no additional codes and themes were robustly supported by data from participants. The study recruited 14 participants to reach saturation.

### Data analysis

Thematic analysis was employed using an inductive approach to systematically identify, organize, and interpret patterns within the interview data. This process followed the six-step framework outlined by Braun and Clarke (23), which involved familiarization with the data, generating initial codes, searching for themes, reviewing themes, defining and naming themes, and writing the report. This method was suitable for identifying, analysing, and reporting patterns within the data, allowing for a detailed and nuanced understanding of the participants’ experiences and perspectives (23). The researchers began by transcribing and thoroughly familiarizing themselves with the interview data, allowing them to immerse themselves in the content. Next, significant features of the data were systematically coded, capturing key elements relevant to the research questions. The initial codes were organized into broader themes, which were subsequently refined and broken down into subthemes to enhance analytic depth and capture variation within participants’ experiences. These themes underwent an iterative and rigorous review to ensure coherence, distinctiveness, and close grounding in the data, after which each theme was clearly defined and named to encapsulate its conceptual essence. Finally, the researchers integrated these themes into a cohesive narrative, using participants’ quotes to illustrate key points and provide a rich, detailed account of how gynecological cancers and their treatments affect fertility among women receiving care at the Ho Teaching Hospital in Ghana. At the end of each interview, participants were invited to clarify or add any further information.

### Rigor

To ensure the trustworthiness of the study, the key quality criteria outlined by Kumar et al. (24) were carefully followed. Confirmability was pursued through strategies such as prolonged engagement, reflexivity, independent coding, and peer evaluation. Transferability was strengthened by providing rich descriptions of participants’ characteristics, their accounts of the phenomenon, and the researcher’s contextual observations, in line with Polit and Beck’s (25) guidance. Dependability was addressed by conducting a systematic review of transcripts and engaging in collaborative discussions with team members to validate coding. Credibility was enhanced by using member-checking techniques, such as playing back interviews to participants for clarification and confirmation of their perspectives. Collectively, these measures established a strong foundation of trustworthiness in the qualitative research methodology of this study.

### Ethical considerations

Ethical approval for this study was obtained from the University of Health and Allied Sciences Research Ethics Committee (Approval No. UR199/1125). Ethical approval for the study was from October 2025–October 2026. All research activities were conducted in accordance with established ethical principles, including respect for persons, privacy, confidentiality, beneficence, and non-maleficence, to safeguard the rights, dignity, and well-being of participants throughout the study. Written informed consent was obtained from all participants prior to data collection. Participants were provided with detailed information regarding the study’s purpose, procedures, potential risks, and benefits, and were assured that participation was entirely voluntary. Interviews were scheduled at times and conducted in private locations that ensured comfort and confidentiality. Participants were informed of their unrestricted right to withdraw from the study at any point without penalty or any effect on their clinical care. Confidentiality was rigorously maintained by anonymizing all data and removing identifying information from transcripts, records, and reports. Audio recordings and transcripts were securely stored and accessed only by members of the research team. No personally identifiable information was included in any dissemination of findings.

Given the sensitive nature of discussions surrounding fertility loss and reproductive concerns related to gynecological cancers and their treatment, interviews were conducted with empathy, sensitivity, and cultural awareness. Participants were continuously monitored for signs of emotional distress. When participants became visibly emotional, interviews were paused to provide reassurance and support, and participants were reminded of their right to discontinue participation. Where appropriate, participants were gently referred to the hospital’s psychosocial support services for additional emotional assistance. Beneficence guided the overarching aim of the study, which sought to generate evidence to inform culturally responsive, fertility-sensitive care for women affected by gynecological cancers. Cultural beliefs and norms related to fertility, womanhood, and reproductive decision-making were respectfully acknowledged, even when they differed from biomedical perspectives. Through these ethical safeguards, the study aimed to minimize potential harm while contributing meaningful insights to improve reproductive health care and support for women receiving care at Ho Teaching Hospital in Ghana.

### Findings

#### Sociodemographic characteristics of participants

The study recruited 14 participants to reach saturation. Most participants (6; 42.85%) were aged 20-30 years. The youngest participant was between 21 and 25, while the eldest was between 46 and 50 years old. The majority (9; 64.28%) were single and had no children. Almost half (6; 42.85%) were schooled up to the tertiary level. The majority (10; 71.42%) are from the Ewe tribe. All participants profess the Christian faith.

**Table 1.** Socio-demographic information of participants.

| Participants | Age (Years) | Marital status | Number of children | Type of cancer | Educational level | Occupation | Religion | Ethnicity |
| --- | --- | --- | --- | --- | --- | --- | --- | --- |
| Woman A | 26–30 | single | None | Ovarian | Tertiary | Manager | Christian | Ewe |
| Woman B | 21–25 | single | None | Cervical | SHS | Student | Christian | Ewe |
| Woman C | 26–30 | married | Four | Cervical | Primary | Farmer | Christian | Ewe |
| Woman D | 46–50 | single | None | Uterine | Tertiary | Teacher | Christian | Ewe |
| Woman E | 36–40 | single | None | Ovarian | Tertiary | Teacher | Christian | Ewe |
| Woman F | 36–40 | Single | None | Cervical | SHS | Trader | Christian | Ewe |
| Woman G | 46–50 | Married | Four | Endometrial | Primary | Unemployed | Christian | Ewe |
| Woman H | 36–40 | Married | Two | Ovarian | SHS | Unemployed | Christian | Akan |
| Woman I | 26–30 | Single | None | Cervical | Primary | Trader | Christian | Ewe |
| Woman J | 26–30 | Single | None | Ovarian | JHS | Trader | Christian | Akan |
| Woman K | 31–35 | Married | One | Cervical | SHS | Orderly | Christian | Ewe |
| Woman L | 31–35 | Married | Three | Ovarian | Tertiary | Teacher | Christian | Ewe |
| Woman M | 26–30 | Single | None | Cervical | Tertiary | Student | Christian | Akan |
| Woman N | 21–25 | Single | None | Cervical | Tertiary | Student | Christian | Akan |

#### Presentation of Themes

The themes were developed from patterns identified in participants’ narratives, reflecting the psychological, emotional, social, and cultural dimensions of living with the condition. Each theme is accompanied by subthemes that provide a deeper understanding of the specific aspects within these broader categories. In all, the analysis yielded three themes and thirteen subthemes

**Table 2.**
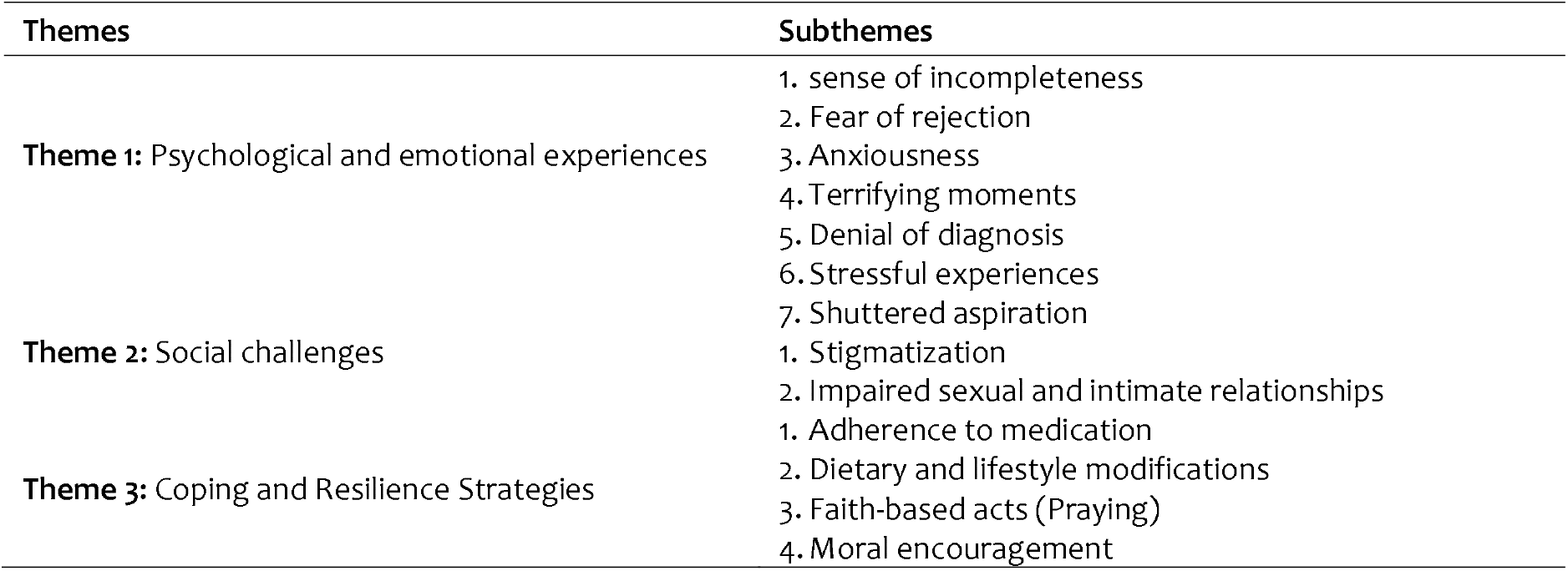
Presentation of emerged Themes and Sub-Themes.

### Theme 1: Psychological and emotional experiences

The study found that most participants faced a range of psychological challenges, including feelings of incompleteness and fear of rejection. The experiences of participants extended beyond the psychological domain into deep emotional involvement, with most reporting feelings of fear, anxiety, denial, stress, and brokenness following their diagnosis. The narratives revealed how gynecological cancer profoundly affected their emotional well-being, shaping their perceptions of life, health, and motherhood.

### Subtheme 1: Sense of incompleteness

A dominant emotional thread expressed by participants was a profound sense of incompleteness stemming from the reality of infertility. In the Ghanaian cultural context, womanhood is deeply intertwined with motherhood, and the ability to conceive is often seen as a core marker of feminine identity and social worth. For many women, the confirmation of infertility triggered feelings of personal inadequacy, social failure, and internal conflict. Participants described a loss that extended beyond biological motherhood by saying:

> *Sometimes I feel incomplete as a woman because of the fear and thought that I may not be able to give birth*. **Woman A**
>
> *At first, I felt incomplete as a woman because I could no longer have children. I used to think that being able to give birth was what made a woman whole, but through this experience, I have learned that womanhood is more than that. It is about strength, care, and love. I now believe that motherhood is not only about giving birth but also about nurturing and caring for others. My view of life and womanhood has changed, I have become stronger and more understanding, and I now value emotional and spiritual strength more than physical ability*. **Woman F**

### Subtheme 2: Fear of rejection

Participants frequently expressed a deep-seated fear of rejection, which significantly shaped their decisions regarding romantic relationships. The diagnosis of infertility created anxiety over potential social stigma and partner abandonment, leading many to avoid intimate relationships altogether. Participants described how their diagnosis reshaped their social lives, prompting them to withdraw from pursuing romantic involvement to avoid anticipated judgment or emotional pain. They expressed this concern by saying:

> *I was not in a romantic relationship before my diagnosis, and after knowing my condition, I have lost interest in entering into one because I am afraid of being rejected due to infertility. I haven’t told anyone else apart from them because I fear being stigmatized or pitied*. **Woman F**
>
> *I didn’t have a partner; I don’t intend having one again. I can’t conceive and most of our men want women who can give them children. […] I need to focus on my education*. **Woman N**

### Subtheme 3: Anxiousness

The uncertainty surrounding prognosis, treatment, and fertility outcomes generated high levels of anxiety among participants. Many reported that the emotional stress of their diagnosis affected both personal decisions and social interactions. They expressed their worries by saying:

> *I was sad and anxious about the future since I knew I won’t be able to conceive anymore. It didn’t affect my relationship with my family (mum and dad) but here were rather supportive and encouraged me to be positive. I was not in a romantic relationship prior to the diagnosis. I do feel it has negatively influenced my decision not to enter into a romantic relationship because of my infertility*. **Woman F**
>
> *I was stressed and worried about my health and life when I was first told about my condition. I was informed that the surgery would make me unable to give birth, but I wasn’t bothered since I already had four children*. **Woman G**
>
> *Emotionally I was stressed and anxious, and I wasn’t stigmatized because nobody knew about my condition*. **Woman E**

### Subtheme 4: Terrifying moments

Participants described being terrified at the moment of diagnosis, often fearing death, the unknown progression of the disease, and social judgment. Many emphasized how confusing and overwhelming the situation felt while trying to stay strong in the face of fear, stating:

> *I went to the hospital, and after several tests, the doctor said there was a tumor on my ovary. I was very scared at first, but I tried to stay strong*.**Woman B**
>
> *I was very scared and confused because I didn’t know much about it. The treatment journey was difficult, with constant checkups and fear of what would happen next*. **Woman E**
>
> *I was afraid of dying and also of what people might say. However, with time, my parents encouraged me and supported me through the treatment. The surgery was painful, but I thank God that I survived it. Living with the condition has taught me to be strong and appreciate life more*. **Woman F**

### Subtheme 5: Denial of diagnosis

Participants reported that after the diagnosis, they initially refused to believe it, struggling to accept that they had cancer. The disbelief was often coupled with worry and emotional turmoil, making it difficult to process the news. They expressed their disbelief by saying:

> *I decided to go to the hospital, and after several tests, I was told I had uterine cancer. Initially, I didn’t accept the diagnosis when the doctor informed me. But later, I had to accept it, though very difficult. I became very emotional and worried afterwards about what would happen to me*. **Woman B**
>
> *[…] I went to the hospital and after some tests, I was told I had endometrial cancer. When I heard the news, I didn’t believe it. I was shocked, I thought it was just something normal happening to me, not knowing it was cancer. For days, I was denying that it can be true*. **Woman M**

### Subtheme 6: Stressful experiences

Participants expressed how the diagnosis and treatment created ongoing stress, with concerns about surgery, recovery, and the uncertainty of the future. Many described feeling emotionally drained and overwhelmed by the challenges they faced. They described this period as one of the most difficult times in their lives.

> *The journey after I was informed about my condition was extremely stressful. I was constantly worried and overwhelmed, especially when I thought about the surgery and the uncertainty of what would happen to me. I could not sleep well, and my mind was always racing with fear*. **Woman H**

Stress was often intensified by repeated hospital visits and exposure to other patients’ experiences, which heightened anxiety and fear.

> *Each hospital visit increased my stress. Seeing other patients and hearing their stories made me more afraid*. **Woman G**

For some participants, the prolonged nature of illness and physical decline compounded psychological distress, leading to feelings of despair and emotional breakdown.

> *I cried a lot because I felt my life was over. The past two years have been very hard. I’ve been bedridden and weak most of the time. It is very stressful*. **Woman C**

Others similarly described a sense of constant emotional strain and depletion:

> *I was always thinking, always worrying. Even when nothing was happening, my heart was not at rest*. **Woman D**

### Subtheme 7: Shuttered aspiration

Many participants, particularly those who were single, felt devastated by the potential loss of their ability to conceive. They described sadness, grief, and uncertainty about fulfilling their aspirations of motherhood, while some tried to mentally prepare for the possibility of infertility. They expressed their sorrow by saying:

> *Even after treatment, they couldn’t guarantee that I would be able to get pregnant. Hearing this broke my heart because I realized I could not have the children I had always dreamed of having*. **Woman A**
>
> *The doctor said my womb was not removed, so I could still conceive, but at my age and after the surgery, I’m not sure if it’s possible. That thought sometimes makes me sad because I’ve always wished to be a mother*. **Woman D**
>
> *Before the surgery, the doctor explained that they would have to remove my uterus, and that meant I would not be able to give birth again. Hearing that was very painful for me because I had always hoped to have children someday*.

It is worth noting that participants who already had more than one child were not particularly disturbed by the news that they would no longer be able to conceive. One participant remarked:

> *The surgery and treatment affected my fertility, and I can no longer conceive. However, I accepted it peacefully because I already have children and felt content with that. Yes, I was told that the surgery would make me unable to give birth. I wasn’t worried or disturbed since I already had four children and was not planning to have more*. **Woman G**

### Theme 2: Social challenges

Women diagnosed with gynecological cancer often experience profound social challenges that extend beyond their physical illness. Participants highlighted that their diagnosis and subsequent infertility had a significant impact on how they were perceived in their communities, affecting their relationships, social standing, and daily interactions. The social repercussions of the disease were compounded by a general lack of awareness about gynecological cancers, which led to misconceptions and judgmental attitudes.

### Subtheme 1: Stigmatization

Participants described being stigmatized by community members, colleagues, and even friends who lacked understanding of their condition. In some cases, others falsely assumed they had HIV/AIDS or labeled them as barren simply because they could not conceive after treatment. This stigmatization caused participants to feel alienated, embarrassed, and emotionally vulnerable.

> *People have been stigmatizing me and some even say I’ve gotten HIV/AIDS. Some also say I can’t give birth again*. **Woman C**
>
> *I’ve been stigmatized by colleagues and friends in conversations. But usually, they don’t say it to me directly, but I know I’m the one they’re talking about. They don’t know I have this condition, but they know I don’t have children, so they think I’m barren*. **Woman D**

### Subtheme 2: Impaired sexual and intimate relationships

The participants also discussed how the physical and physiological effects of gynecological cancer disrupted intimate relationships, particularly with their spouses. The condition affected their reproductive organs, causing symptoms such as bleeding, discharge, and pain that made sexual intimacy challenging or impossible. These changes not only caused physical discomfort but also emotional strain, as women felt unable to fulfill what they considered expected marital roles. They expressed their frustration by saying:

> *Since I was diagnosed, I haven’t been intimate with my husband, which makes my husband worry sometimes*. **Woman K**
>
> *Before the condition, I was a married woman, and still I am. But due to the condition, I can’t fulfill all my obligations as a woman, there are times my husband will be in need of me as a woman, but due to my condition, there is nothing we can do*. **Woman G**

### Theme 3: Coping Strategies

This theme highlights strategies participants employed to cope with their diagnoses and the challenges associated with treatment. Participants demonstrated proactive engagement with their healthcare plans, emphasizing adherence to prescribed treatments, lifestyle modifications, and dietary changes. These coping steps reflect an understanding of the importance of self-management and personal responsibility in improving health outcomes and preventing complications.

### Subtheme 1: Adherence to medication

Participants expressed a strong commitment to following their prescribed treatment regimens, including taking medications on schedule and undergoing surgeries as recommended by healthcare professionals. This adherence was motivated by a desire to recover and regain control over their lives. They expressed this commitment by saying:

> *This was a condition I knew nothing about. The most thing that helped me is that I paid attention to what the doctor said. I bought the medications they prescribed and took them as ordered. Also, the surgery I did helped me in a way that it gave me hope that I will be fine*. **Woman B**
>
> *Personally, I just focus on taking my medication regularly as I was taught by the healthcare workers*. **Woman A**

### Subtheme 2: Dietary and lifestyle modifications

Participants reported making intentional dietary changes to support their treatment, promote recovery, and prevent the progression of their condition. Many had adjusted their eating habits by reducing foods perceived to exacerbate their illness and increasing the consumption of nutritious, health-promoting foods.

> *I used to take carbonated drinks, but after I was diagnosed with cancer, I did my own research, and I found out that the carbonated drinks are not good for someone with cancer. So, I’ve stopped consuming them totally. I also eat healthy nowadays with little consumption of carbohydrates and more of vegetables, fruits, and water*. **Woman D**
>
> *I also adjusted my lifestyle towards healthy eating and doctors advised me against too much consumption of salt*. **Woman B**
>
> *As for me I don’t eat anyhow like taking of these carbonated drinks or smoked fish is a big no for me. So, after the diagnosis I stuck strictly to a healthy diet. I added more vegetables to my food*. **Woman E**

### Subtheme 3: Faith-based acts

All participants identified as Christians and emphasized turning to prayer and spiritual practices as a source of strength and healing. Faith served as both a coping mechanism and a source of emotional support, helping participants navigate the uncertainty and fear associated with their diagnosis. They expressed their faith by saying:

> *I am a Christian, and I know God is the ultimate healer. So, I always prayed for divine healing because without God wouldn’t be alive*. **Woman B**
>
> *I prayed for mercy from God. Before the surgery, the pastor came to pray for me. God hears me anytime I pray, He is still healing me*. **Woman H**
>
> *I normally pray to God to strengthen me mentally and emotionally so that it doesn’t weigh me down and I feel He has indeed strengthened me*. **Woman F**

### Subtheme 4: Psychological Support

Participants also drew strength from the encouragement and support of family members and close friends. This moral support helped them maintain a positive outlook and reinforced their motivation to adhere to treatments and pursue recovery. One participant explained:

> *I also receive encouragement from my parents and siblings and that has also helped me a lot in not dwelling on my condition but I feel my mom has helped me the most in dealing and accepting my condition. After my surgery the doctor also said I can adopt a child if I feel the need for one*. **Woman F**

Another participant highlighted the support of a spouse and family members with healthcare knowledge:

> *My husband encourages me a lot and tells me that things will get better with time. My sister’s daughter, who is a nurse, have supported me the most* **Woman G**
>
> *[…] My parents comforted me and reminded me that life is more important and that I can still be a mother in other ways, such as adoption*. **Woman J**

## Discussion

Gynecological cancers and their treatments profoundly disrupt fertility at a stage when reproductive potential is central to women’s identities, social positioning, and future life plans (1). In exploring this impact among women of reproductive age at Ho Teaching Hospital, Ghana, this study reveals fertility loss as a complex and enduring consequence of cancer that extends well beyond physiological impairment. Participants’ accounts illuminate deep social and psychological stressors embedded within gendered and cultural expectations of womanhood and motherhood. These experiences were further compounded by treatment-related morbidity, stigma surrounding reproductive cancers, financial strain, and disruptions to education, collectively intensifying emotional distress and shaping how women navigated diagnosis, treatment, and survivorship. Despite these challenges, participants articulated adaptive coping strategies grounded in treatment adherence, faith and spirituality, and social support, which fostered resilience and sustained hope. The findings underscore the need for holistic, culturally responsive care that integrates fertility counseling, psychosocial support, effective symptom management, and social protection for women living with gynecological cancers.

Participants consistently described fertility impairment as a profound disruption to self-concept, often articulated through feelings of incompleteness, diminished womanhood, and loss of purpose. This feeling is not merely biological but socially constructed, especially in contexts like Ghana, where motherhood is strongly tied to social acceptance and respect. Prior evidence emphasizes that infertility challenges women’s sense of self and belonging, often resulting in diminished self-worth (26). Consistently, cancer survivors who experience infertility describe feelings of being “incomplete” or “broken,” highlighting the deep emotional toll of fertility loss (27,28).

Fear of rejection further intensified sources of distress, reflecting marital and gender norms that prioritize childbearing and position infertility as a relational risk (29). Anticipated abandonment and social marginalization due to infertility undermined women’s self-worth and discouraged engagement in intimate relationships, amplifying emotional vulnerability. Similar findings were noted in a study revealing that women with fertility issues often feared that their partners may leave them for more “fertile” women (30). Against this backdrop of social and relational insecurity, diagnosis and treatment decision-making were frequently described as terrifying moments marked by existential fear, uncertainty, and anticipatory grief, not only for threatened survival but also for foreclosed motherhood. These intersecting threats intensified cancer-related distress and shaped how women interpreted, navigated, and coped with treatment, particularly when survival was framed as coming at the expense of reproductive potential (31).

Stigmatization was a significant social stressor experienced by women living with gynecological cancers, arising from deeply rooted misconceptions that associate illness affecting reproductive organs with moral, sexual, or spiritual transgression (32). Such stigma not only precipitated social isolation and judgment but also reinforced gendered expectations surrounding purity, fertility, and womanhood. These findings are consistent with a study that reported that stigma surrounding reproductive cancers remains a significant barrier to early diagnosis and support, often leaving women feeling alienated and emotionally distressed (33). In a similar vein, evidence showed that women with cervical or ovarian cancer often experienced feelings of embarrassment and self-blame, which hindered their willingness to seek help or disclose their condition (6). The resulting avoidance and social isolation experienced by participants eroded social support networks, intensified psychological distress, and discouraged open disclosure and timely engagement with care, amplifying the overall burden of the disease beyond its physical manifestations.

The findings revealed that women’s coping strategies were deeply shaped by both medical knowledge and sociocultural values, illustrating adaptive efforts to regain control and sustain hope amid illness. Adherence to prescribed medications and recommended nutritional practices functioned not only as treatment compliance but also as psychologically empowering actions that reinforced agency, trust in healthcare providers, and optimism about recovery (34–36). Alongside these biomedical strategies, faith and spirituality emerged as central emotional resources, with prayer providing comfort, reducing anxiety, and offering existential meaning in the face of uncertainty (37,38). Moral encouragement from family and social networks further strengthened resilience by mitigating isolation, enhancing emotional stability, and reinforcing adherence to treatment. This underscores the critical role of social support in sustaining coping capacity and psychological well-being throughout the cancer trajectory.

## Limitations

Despite the important insights generated, this study has several limitations. First, the study relied on participants’ subjective accounts of their fertility experiences and treatment-related impacts, which may be influenced by recall difficulties, emotional distress, or social desirability, potentially shaping how women described sensitive issues such as infertility, stigma, and sexual functioning. Second, the study was conducted in a single tertiary referral facility (Ho Teaching Hospital), and participants were purposively recruited; therefore, the experiences captured may not fully reflect those of women receiving care in other regions of Ghana, lower-level facilities, or women who disengaged from care. Third, the qualitative descriptive design enabled an in-depth understanding, but does not allow quantification of the magnitude of fertility impairment or comparison of experiences by cancer type, treatment modality, or time since treatment, and causal inferences cannot be made. Finally, because women who were critically ill or with mental health concerns were excluded, the study may under-represent the experiences of those with the most severe psychosocial burden.

## Conclusion

The study revealed that women whose fertility has been affected by gynecological cancers experience profound psychological, emotional, social, and economic challenges that deeply shape their quality of life. Feelings of incompleteness, fear of rejection, stigma, and financial strain were central to their lived realities. Despite these hardships, many women demonstrated resilience through faith, moral encouragement, and adherence to medical advice. These findings underscore the need for a holistic and culturally sensitive approach to care, one that recognizes the emotional and spiritual dimensions of healing alongside clinical management. Strengthening supportive care systems, improving public awareness, and enhancing the role of nurses in psychosocial interventions will be essential in promoting recovery and dignity among women living with gynecological cancers.

## Author contributions

A.A. and D.B. A. conceived the study and led the data analysis and manuscript drafting. F.C., P.A., O.A.G., M.V., B.A., R.A.A., M.A.A., S.S.D., D.B.D., S.M.S., conducted the literature search and contributed to writing the manuscript. All authors critically reviewed the manuscript for important intellectual content, contributed revisions, and approved the final version for submission.

## Data availability

The datasets generated and/or analyzed during the current study are not publicly available due to the sensitive and confidential nature of the participants’ personal and medical information, as well as ethical restrictions imposed by the institutional review board. However, de-identified data may be available from the corresponding author on reasonable request and subject to approval by the ethics committee.

## Declarations

### Ethical approval and consent to participate

The researchers sought ethical review and clearance from the Research Ethics Committee of the University of Health and Allied Sciences before commencing data collection (Approval No. UR199/1125). Written informed consent to participate was obtained from all of the participants in the study. The study followed the principles and guidelines of the Declaration of Helsinki.

### Consent to publish

Not applicable.

## Acknowledgment

Not applicable.

## Appendix

### S1 Appendix: Interview Guide

#### A: Background Information

1. Can you please tell me a little about yourself?
  a. age
  b. marital status
  c. number of children
  d. occupation
  e. educational level
  f. religion
  g. type of cancer
  h. ethnicity

#### B: Perceptions and Understanding of Gynecological Cancers and Fertility

1. What do you know about gynecological cancers?
  a. Probe: How did you come to learn about it? (e.g., media, health professionals, family/friends)
2. What is your understanding of how gynecological cancers may affect fertility?
  a. Probe: Do you think it can influence a woman’s ability to have children? How?
3. What concerns, if any, do you have about fertility and gynecological cancers?

#### C: Challenges Faced by Women with Gynecological Cancers

1. Since your diagnosis, what challenges have you faced in your day-to-day life?
  a. Probe: Physical challenges (pain, treatment side effects, infertility, etc.)
  b. Probe: Emotional/psychological challenges (stress, anxiety, stigma, relationships)
  c. Probe: Financial/social challenges (cost of treatment, support from family/community)
2. How have these challenges affected your family life, work, or social relationships?
3. Are there specific cultural or community-related challenges you have experienced?
4. Have you experienced any form of stigma or discrimination because of your condition?
  a. Probe: From family members (changes in how they relate to you)
  b. Probe: From friends or colleagues (exclusion, labeling, gossip, reduced support)
  c. Probe: From the wider community (social isolation, negative cultural beliefs, misconceptions about cancer)

#### D: Coping Strategies

1. How have you been coping with the challenges you just described?
  a. Probe: What practical steps do you take (e.g., lifestyle adjustments, seeking medical support)?
  b. Probe: What emotional or psychological strategies do you use (e.g., prayer, counseling, support groups)?
2. Who or what has supported you most in dealing with your condition?
  a. Probe: Family, friends, religious groups, health professionals, community networks.
3. What advice would you give to other women diagnosed with gynecological cancers about coping with challenges?

#### E: Conclusion

1. Is there anything else you would like to share about your experience with gynecological cancer that we have not discussed?
2. What do you think health professionals or policymakers could do to better support women living with gynecological cancers?

Thank you

